# Spatial Clustering and Multivariate Typologies during lifespan of Hospitalized Traumatic Brain Injury Cases: A Population-Based Study

**DOI:** 10.1101/2025.08.25.25334340

**Authors:** A. Ayma, Q. Legrand, N. Fellay, G. Kathari, D. Delannoy, A. Nouri, E. Beanato, R. Ronchi, K. Schaller, S. Joost, G. R. Iannotti, P. Voruz

## Abstract

**Introduction:** This study examines the demographic, temporal, and spatial patterns of traumatic brain injury (TBI) hospitalizations in the canton of Geneva, Switzerland, between 2012 and 2024. Particular attention is given to differences between seniors (≥60 years) and the younger or working-age population (15–59 years).

**Methods:** Hospital discharge data were analyzed in combination with geographic information systems (GIS) to explore incidence, severity, and clinical outcomes. Temporal trends were assessed across seasons and days of the week. Spatial distribution was investigated using Join Count and SPARR methods to detect clustering. Multivariate clustering further integrated socio-clinical characteristics, including age, autonomy, residential setting, and hospital trajectory.

**Results:** Seniors accounted for more than half of all TBI hospitalizations. Advanced age and institutional residence were strongly associated with reduced likelihood of returning home post-discharge (statistical values). Temporal analyses showed a marked increase in TBI incidence among seniors during autumn and at the beginning of the week, patterns not observed in the working-age group. Spatial analyses revealed non-random clustering of cases, with high-risk zones concentrated in specific areas, particularly for older adults. Multivariate clustering identified distinct socio-clinical profiles in both age groups, shaped by residential environment, autonomy level, and care pathways.

**Conclusion:** The findings underscore the dual clinical and territorial dimensions of TBI, highlighting significant age-related and spatial disparities. Tailored prevention strategies, safer environmental design, and integrated care pathways are needed to reduce inequalities and improve outcomes. Addressing TBI through both a medical and public health lens is essential to support at-risk populations, particularly seniors.

## Introduction

Traumatic brain injury (TBI) is a leading global public health concern, affecting millions of individuals each year and resulting in significant human, social, and economic consequences ^1^. Furthermore, rates of TBI have been increasing, particularly amongst elderly, likely due to the aging population in developed countries. It is defined as an alteration in brain function or evidence of brain pathology caused by an external force, such as an impact, jolt, or penetration. In Switzerland, over 20,000 individuals are hospitalized annually due to TBI ^2^ generating an estimated cost of 300 million Swiss francs for accident insurance systems. These costs are likely underestimated, as they account only for direct medical and rehabilitation expenses, excluding indirect burdens such as productivity loss, caregiver strain, reduced quality of life, and the broader social and economic impacts ^3^.

TBI affects individuals across all age groups, but patterns of risk and outcomes vary significantly between populations. In high income countries, falls have now overtaken road traffic accidents as the leading cause of TBI, although vehicular collisions remain a significant contributor ^1^. Indeed, among seniors, falls are the predominant cause, with both incidence and severity rising significantly after age 65 ^6^. Older patients face unique clinical challenges, including slower recovery, greater risk of complications, and higher mortality rates, often linked to polypharmacy, comorbidities, and the use of anticoagulants ^5^. Moderate to severe TBIs in seniors demand systematic hospitalization, whereas mild cases often go unreported, contributing to gaps in surveillance data ^3^.

However, TBIs are also a major cause of morbidity and mortality among working-age adults. The outcomes of moderate to severe TBI in this group are highly variable, influenced by factors including the severity of actual injury, such as nature of the trauma (i.e. penetrating injury, blunt trauma), the degree of parenchymal injury, and whether surgical intervention is required, as well as non-injury related factors, such as cognitive reserve (including educational level, and social stability^4,5^. While it seems clear that an increase in severity will have worse outcomes, it has also been shown that individuals with higher cognitive reserve tend to achieve better functional recovery and reintegration into working-age life.

While individual-level risk factors (age, sex, medical history) have been well studied across both populations, the influence of contextual factors, such as socioeconomic status, urban environment, and geographic location has received comparatively less attention. Emerging research suggests that the spatial distribution of TBIs is not random but influenced by neighborhood-level deprivation, population density, and disparities in access to emergency and rehabilitation services ^7^. Spatial clustering of cases and geographic disparities point to the need for integrated geospatial approaches in TBI research.

In this context, Geographic Information Systems (GIS) and spatial analysis tools offer powerful means to identify high-risk areas, link injury incidence to local demographic profiles, and support targeted public health interventions. For example, studies in Nova Scotia have identified spatial hotspots of TBI and associated them with economic vulnerability and healthcare access disparities ^7^. However, few studies have applied advanced spatial methods to distinguish risk factors between subpopulations, and none have conducted fine-grained spatial analyses of moderate to severe TBI among seniors in the Swiss context.

This study aims to fill that gap by examining the spatial distribution of moderate to severe TBIs in the canton of Geneva between 2012 and 2024 (*n* = 1071), with a focus on both the working-age (< 60 years) and elderly populations (> 60 years), based on the World Health Organization’s (WHO) definition of older adults for epidemiological purposes ^8,9^. We hypothesize that TBI incidence in both groups is spatially heterogeneous but linked to distinct sets of clinical, socioeconomic, and environmental factors. Moreover, we hypothesized a temporally structured relationship between TBI incidence and time-related factors, namely season and weekday, given that certain days (e.g., weekends) may promote risk-taking behaviors, and that winter conditions may further contribute to such patterns.

To test our hypotheses, we utilized two complementary data sources: hospital records of patients presenting with moderate to severe TBI between 2002 and 2024, and high-resolution sociodemographic data from the Swiss Federal Statistical Office. Using a combination of statistical and spatial analysis, we aimed to identify high-risk zones, profile vulnerable populations, and provide actionable insights for - public health strategies tailored to the specific needs of Geneva’s diverse communities.

## Methodology

This study relied on three main and spatially harmonized data sources to enable an integrated analysis of TBI in the canton of Geneva. Clinical, demographic, and socioeconomic data were combined and analyzed using descriptive statistics, spatial epidemiology tools, and multivariate methods.

### Ethics

This study is based on retrospective data from the Geneva University Hospitals (HUG), collected under the general consent provided by patients, and was approved by the Cantonal Research Ethics Commission of Geneva (CCER: 2025-00737).

### Data availability

The data supporting the findings of this study are available from the corresponding author upon reasonable request.

### Patients and control population

Clinical data were obtained retrospectively from the neurosurgery department records (between 2012 and 2023) of the Geneva University Hospitals (HUG) and included 1’071 patients diagnosed with TBI between 2012 and 2024 who have given a general consent for research. Of these 678 were seniors aged 60 years or older and 393 were working-age adults or (between 18 and 59 years old). Although the official retirement age in Switzerland is 65 years, we selected 60 years as the age cut-off based on the WHO definition of older adults for epidemiological purposes, allowing for early identification of age-related vulnerability in this population ^9^.

For each patient, the dataset included residential address, date of injury, sex, age, primary diagnosis (ICD-10 coded), and post-discharge destination. Residential addresses were georeferenced and projected into the Swiss national coordinate system CH1903+/LV95 (EPSG:2056). Quality control procedures were applied to correct ambiguous or erroneous geolocations. Cases occurring outside the canton of Geneva or Switzerland were excluded. Recurrent injuries were retained in the dataset, as the literature highlights their importance in identifying populations at increased risk of reinjury or underlying vulnerability (Maas et al., 2017).

To compare the population hospitalized for TBI with the general population, a synthetic control population was created using geographic data from the Système d’information du territoire genevois (SITG) which is provided by the Cantonal Office for Population and Migration (OCPM) at a sub-sector scale in order to aggregate the census of the Swiss and foreign population at an intermediate level between the parcel and the municipality, specifically the OCS_POPULATION_SSECTEUR layer which provided detailed demographic data at the statistical sub-sector level. Each sub-sector was normalized based on its total population using the following formula: *POP_norm = round(POP × 10,000 /* ∑*POP)*. This normalization resulted in a total simulated population of 10,000 individuals, as previously validate within spatial analysis ^10^. Random points were then generated in QGIS (“Random points in polygon” function) within the polygons representing each sub-sector. The number of points per sub-sector was proportional to the normalized population. Each synthetic individual was assigned an age and sex based on the demographic distributions observed within their respective sub-sector. Random sampling was performed using a Python function, which first selected an age group proportionally to its frequency, then randomly assigned an age within that group.

### Socioeconomic and Demographic Data

Socioeconomic and demographic indicators were derived from the Swiss Federal Statistical Office (FSO, 2022) and compiled by MicroGIS (2018). Variables included the proportion of residents with tertiary education, the proportion of foreign nationals, men, retirees, and working-age workers, the synthetic overcrowding index (defined as the unweighted mean of person-per-dwelling ratios across six room size categories, from 1 to 6+ rooms, reflecting average occupancy intensity per category), and a deprivation index. This deprivation index was calculated as the difference (in thousands of CHF) between the median income of private households and 60% of the gross median income in the cantons of Geneva. A negative score indicated economic deprivation, while a positive score reflected a standard of living above the deprivation threshold.

Analyses were performed at two spatial resolutions: the statistical sub-sector and the inhabited hectare (100 m × 100 m grid). The hectare level was chosen as a compromise between individual-level precision and the broader aggregation of sub-sectors, which may obscure local heterogeneity. Where data were not available at the hectare level, both spatial scales were used complementarily. Socioeconomic variables were assigned to patients based on their residential hectare, following best practices in environmental epidemiology ^11^.

### Statistical analysis

#### Descriptive Statistical Analysis

An initial descriptive analysis was conducted to explore the distribution of TBI cases by age, sex, and hospital length of stay. TBI incidence rates per 100,000 population were calculated for five-year age intervals using population data from the SITG (OCS_POPULATION_SSECTEUR layer). An age pyramid was produced to compare the demographic profile of TBI cases to that of the general population. Statistical tests were applied to assess group differences: the Mann–Whitney U test for quantitative variables, the χ² test for categorical variables, and the Kolmogorov–Smirnov test to assess distribution normality and justify the use of non-parametric methods.

#### Spatial Relative Risk Analysis

To assess spatial patterns of TBI risk, a control population was simulated using random point generation in QGIS (“Random points in polygon” function). A case-to-control ratio of approximately 1:10 was maintained. Each random point was assigned an age group and sex according to the local distributions observed in the case data, ensuring representativeness. This simulation approach follows recognized best practices in spatial public health analysis ^10^. For subgroup analyses, the procedure was adapted to generate control populations of 6,000 and 4,000 seniors, respectively.

Relative risk surfaces were computed using the Spatial and Spatiotemporal Relative Risk (SPARR) method via the sparr package in R ^10^, with adapted bandwith ^12^. The method compares the spatial density of observed TBI cases to that of the control population using adaptive kernel estimation. The statistical significance of high- and low-risk areas was assessed through Monte Carlo permutations (999 iterations), with *p* < .05 and *p* < .01 used as significance thresholds.

#### Spatial Structure Detection

To further evaluate spatial clustering, the Local Join Count test was performed using GeoDa ^13^. An adaptive bandwidth distance matrix defined the spatial weights (19 for senior and 17 for working-age), based on the default values proposed by GEODA (Kernel function uniform,, diaonal weights=1, distance metric Euclidian). This test determines whether cases are more frequently adjacent to each other than would be expected by chance. The number of black–black (BB) joins was compared against random distributions generated via permutation to assess statistical significance of clustering.

#### Multivariate Analysis and Cluster Profiling

A Principal Component Analysis (PCA) was conducted in GeoDa to reduce dimensionality across individual and contextual variables. The following indicators were included: age, sex, hospital stay duration, return to home (yes/no), long-term care residence (yes/no), deprivation index, overcrowding index, tertiary education rate, proportions of retirees, men, foreign nationals, and working-age workers. All variables were standardized using Z-score transformation to ensure comparability.

The resulting principal components were used for unsupervised classification through Hierarchical Ascending Clustering (HAC). Ward’s linkage method was employed, minimizing intra-cluster variance to produce homogenous groups. The optimal number of clusters was determined through silhouette coefficient analysis and dendrogram inspection.

All results, including relative risk maps, cluster structures, and sociodemographic profiles were visualized using QGIS 3.28. These spatial outputs were used to identify geographic disparities and formulate public health recommendations targeting high-risk senior populations.

The following tools and software were used: QGIS 3.28: Geocoding, random point generation, spatial joins, mapping; GeoDa 1.20: PCA, HAC, Join Count test; R (sparr, sf, spatstat): Kernel-based relative risk estimation; Python: Initial data exploration and statistical testing; GISy: Spatial integration and harmonization tools.

## Results

### Age and Sex Distribution of TBI Patients

The analysis of age distribution among patients hospitalized for TBI, normalized over 100’000 cases for comparison purposes, revealed that TBI incidence increases with age, suggesting a growing vulnerability among older populations (see Figure 1A). Moreover, when compared with standard (synthetic) population, stratified by gender predominance of males was observed, across nearly all age groups (see Figure 1B). Specifically, a chi-square test, assuming no association between sex and TBI occurrence, indicated a statistically significant association between sex and TBI incidence across all age groups except the 55–59 age group, while all other analysis revealed higher incidence rates in men (see Figure 1B).

**Figure 1.**
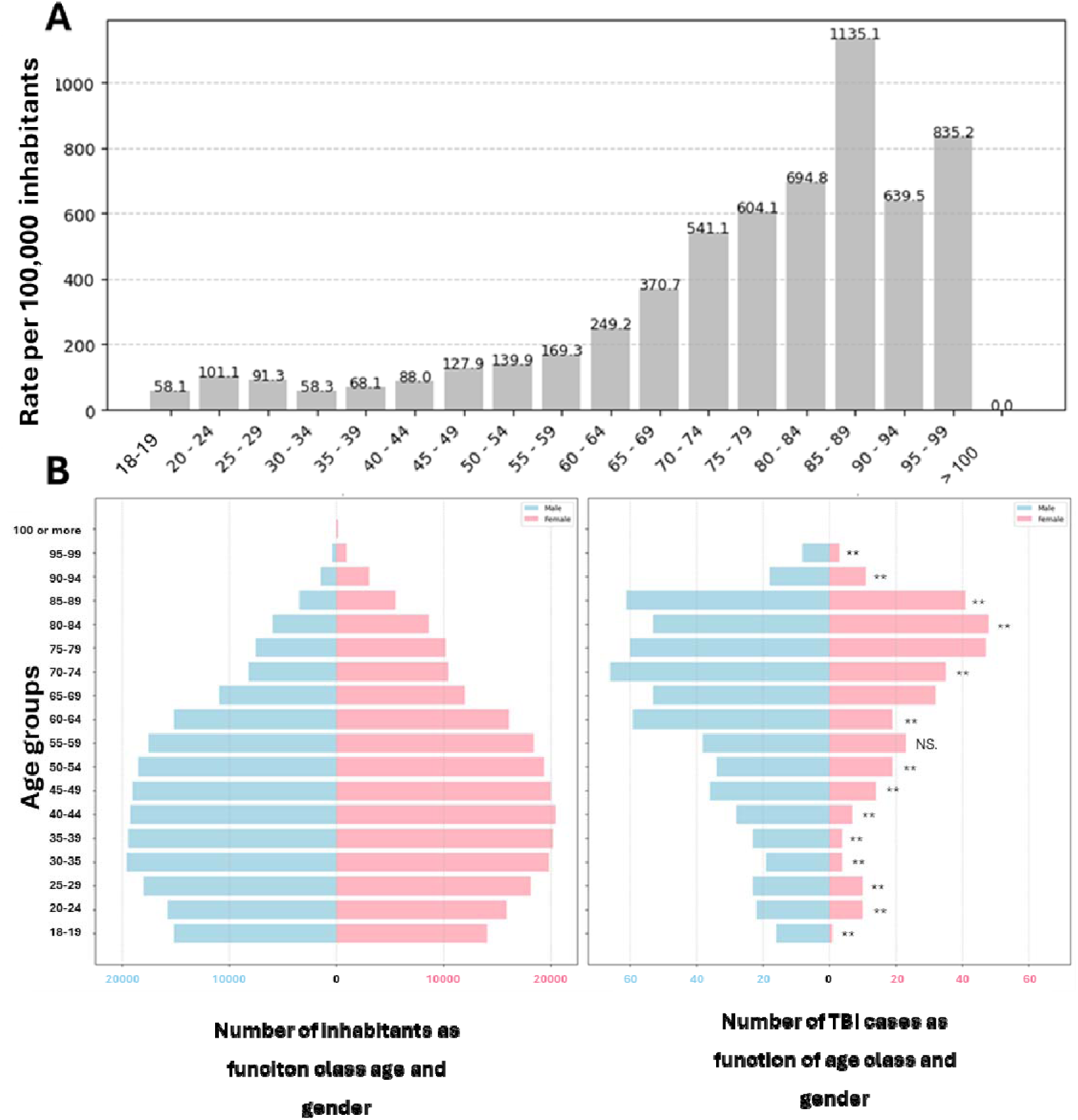
A) Traumatic Brain Injury Incidence by Age Group and as function of sex (normalized per 100,000 inhabitants). B) Swiss Population Age Pyramid (left) and Distribution of Traumatic Brain Injury Clinical Cases (right) by Age and Sex. Bars represent the number of individuals by sex: blue = men, pink = women. While the population pyramid reflects the general age and sex structure in Switzerland, the clinical case distribution reveals a marked overrepresentation of men in nearly all age groups. The sex difference is statistically significant across all age categories, except for the 55–59 age group, where no significant difference was observed.

### Length of Hospital Stay

The analysis of hospital stay duration following TBI on all clinical cases showed a non-normal distribution of the data, as confirmed by the Kolmogorov–Smirnov test (*p* < .050) and therefore Mann–Whitney U tests were conducted to assess differences across age groups. Significant differences in hospital stay duration were observed in the 50–54 and 70–74 age groups for women, and in the 55–59 and 80–84 groups for men. In these cases, individuals of working age tended to have shorter hospital stays than those outside this group.

Additionally, patients aged 15–19 years exhibited significantly shorter hospital stays compared to other age groups (*p* < .050), suggesting that age has a modest yet detectable effect on hospitalization duration, particularly for the youngest cohort.

No significant differences in length of stay were found between sexes.

### Clinical outcome after hospitalization

The analysis of transfer rates to rehabilitation facilities following TBI revealed a marked increase with age. Since the data showed deviation from normal distribution, as confirmed by the Kolmogorov–Smirnov test (*p* < .050) and given the binary nature of the variable (home discharge [1] ; no home discharge [0]), chi-square tests were used to compare proportions of home discharge across age groups.

Among male patients under 60 years of age, several significant differences were found in the rates of home discharge between age groups. For example, men aged 40–44 years had a significantly lower home discharge rate (24%) compared to those aged 45–49 (51%), 35–39 (59%), 30–34 (67%), and 15–19 (67%). Other significant disparities were observed, such as between the 20–24 and 30–34 age groups (26% vs. 67%), and between the 20–24 and 15–19 age groups (26% vs. 67%). While analysis on the whole group using a logistic regression model were used to assess the predictors of home discharge, including sex, age, length of stay, and other contextual and sociodemographic indicators. Among all variables, only length of stay was significantly associated with home discharge (β = –0.121, *p* < .001), indicating that a longer hospital stay is linked to a lower likelihood of returning home. Other variables, including sex, age, population density, education rate, retirement ratio, socioeconomic deprivation index, and local proportions of male, foreign-born, and working populations did not show statistically significant associations (*p* > .050). The model yielded a *R²* of 0.21, suggesting moderate explanatory power.

Among older adults, the home discharge rate after TBI varied by sex and age group. Overall, the rates were lower than those observed in younger populations, ranging from 18% to 63%. A general decline was observed, particularly among men. Among women, higher discharge rates were noted between ages 60–69 before decreasing in older age groups. Chi-square tests comparing home discharge rates by sex within each age group revealed a statistically significant difference only in the 70–74 age group (*p* = .001), while all other age groups showed p-values > .05. However, comparisons across age groups within the same sex revealed additional significant differences. Among men, significant differences were found between those aged 60–64 and 70–74 (*p* = .023), and between 60–64 and 75–79 (*p* = .012). Among women, a significant difference was observed between the 65–69 and 80–84 age groups (*p* = .045). Moreover, as previously done on the working-age group a separate logistic regression analysis was conducted for the senior population to model the probability of home discharge. The model was significant overall (*p* < .001; *R²* = 0.22). Three variables were significantly associated with home discharge: age (*p* < .001), length of stay (*p* < .001), and residence in a nursing home prior to admission (*p* < .001). Sex was not a significant predictor in this model (*p* = .078).

### Weekday and seasonality

Temporal analysis of hospital admissions revealed several significant patterns in TBI incidence. Among older adults, cases were more frequent at the beginning of the week, with significant differences observed between Monday and Sunday (*p* = .037), Tuesday and Sunday (*p* = .004), and Tuesday and Saturday (*p* = .047). Seasonally, a significant difference was found between autumn and spring in the senior population (*p* = .002), with a higher incidence in autumn. Among working-age adults, a significant seasonal variation was also noted, with more TBIs occurring in autumn than in winter (see Figure 2).

**Figure 2.**
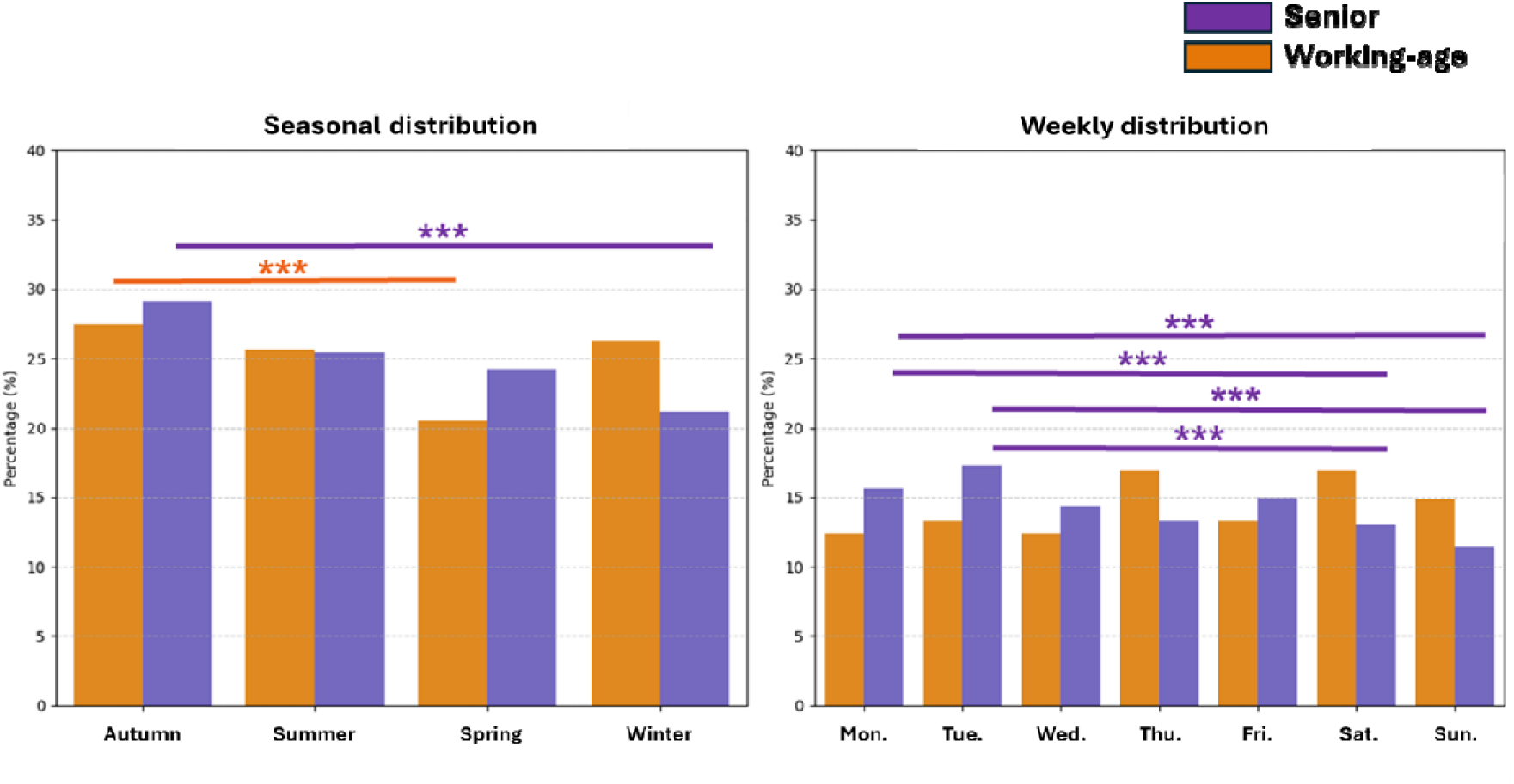
Temporal Distribution of Traumatic Brain Injuries by Age: Comparison of Seasonal and Weekly Patterns Between Seniors and the Working-age Population.

### Spatial distribution of TBI

Spatial analysis using the Join Count and SPARR methods revealed four areas with a statistically significant spatial concentration of TBI cases in the working-age population: the neighborhoods of Geneva–Plainpalais and Carouge, Onex and Bernex, Présinge, and Meyrin(see Figure 4). These findings indicate a non-random clustering of TBI cases in these locations for the working-age population. While the local Join Count test specifically identified several other significant clusters of TBI among older adults, particularly along the border between Cologny and Chêne-Bougeries, as well as in the municipalities of Onex and Veyrier. Additional, though less pronounced, clusters were detected primarily in central Geneva. A high Join Count value combined with a significant *p*-value confirms local aggregation of TBI cases, suggesting that the observed frequency exceeds what would be expected under a random spatial distribution (see Figure 3).

**Figure 3.**
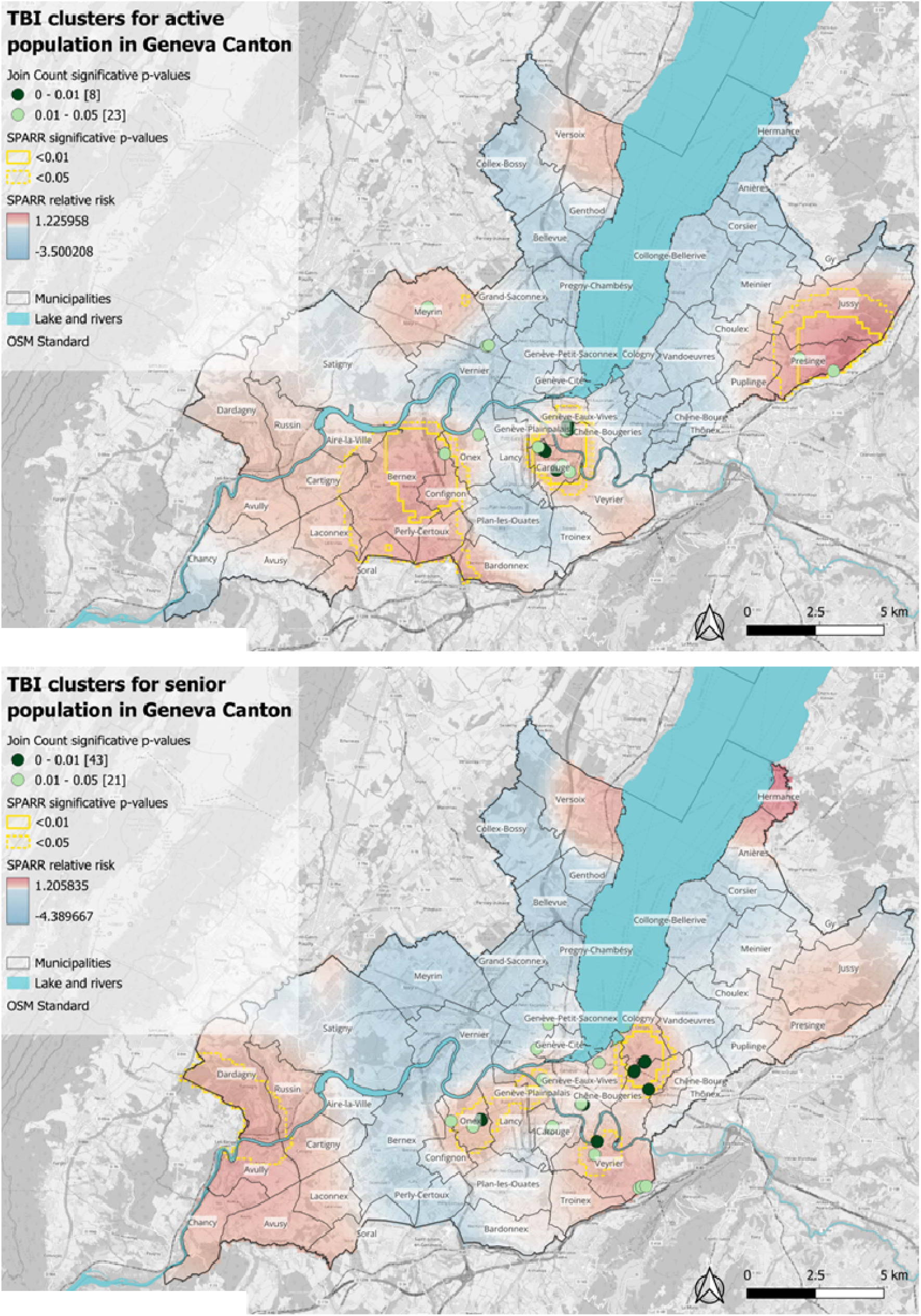
A. Map of Univariate Join Count Clusters and Spatial/Spatiotemporal Relative Risk of Traumatic Brain Injury. B. Mapping of Traumatic Brain Injury Clusters and High-Risk Zones Among Seniors in the Canton of Geneva. Green points indicate spatial clusters identified by the local Join Count test, with light green representing statistically significant clusters (p < 0.05) and dark green highly significant clusters (*p* < .01). Yellow contours delineate areas where spatial relative risk, as estimated by SPARR, is significant (*p* < .05), with darker yellow fill indicating higher significance (*p* < .01). The basemap shows the SPARR-derived relative spatial risk, color-coded from blue (low risk) to red (high risk). Both analyses were performed using an adaptive bandwidth. Municipal boundaries and major waterways are included for geographic reference. Basemap from OpenStreetMap: https://www.openstreetmap.org.

**Figure 4.**
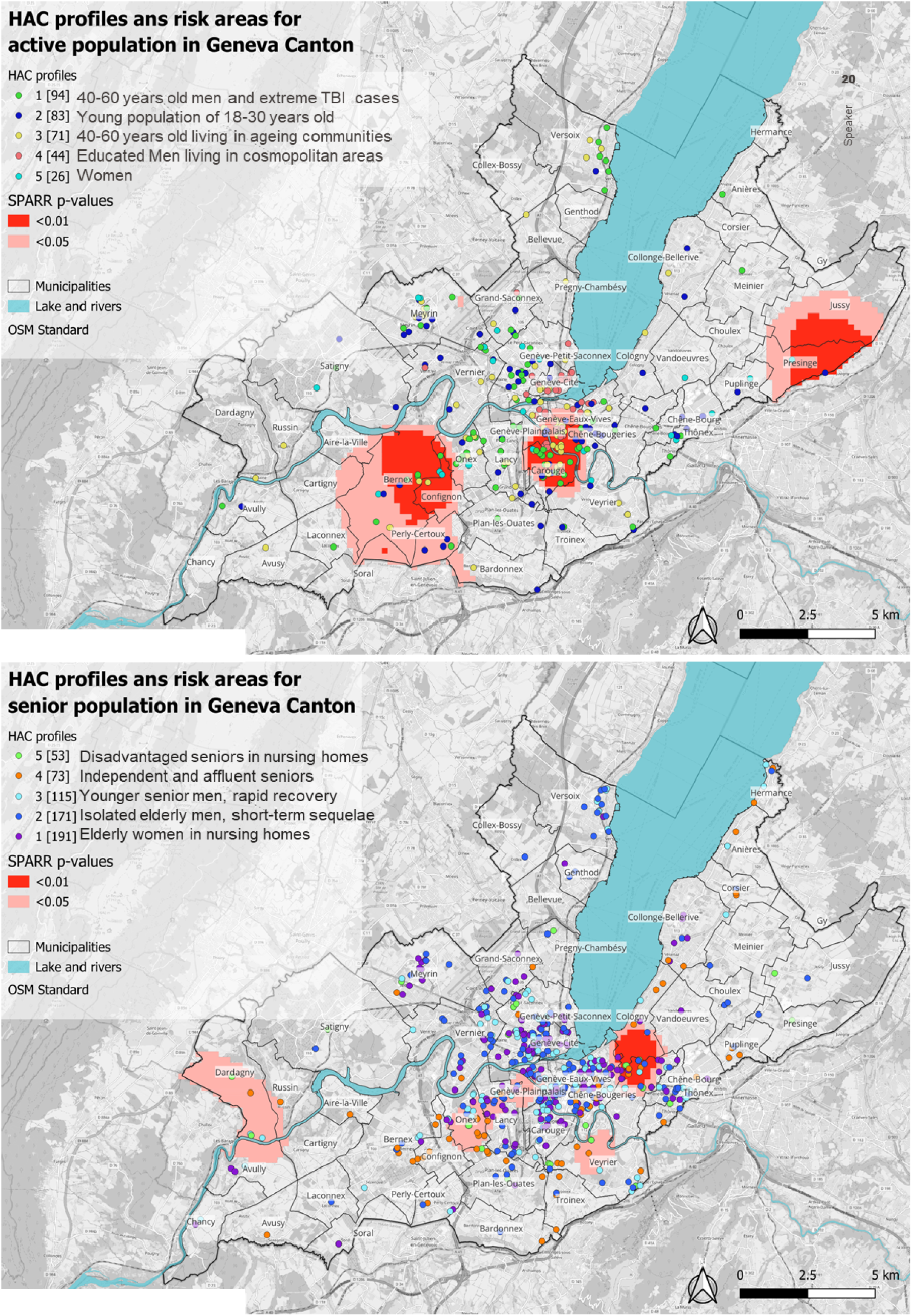
Spatial distribution of working-age and senior clusters. A. Patient profiles identified through hierarchical ascendant clustering (HAC) are represented by colored points: green = Profile 1 (men aged 40–60 with severe TBI cases), dark blue = Profile 2 (young adults aged 18–30), yellow = Profile 3 (adults aged 40–60 living in ageing communities), pink = Profile 4 (educated men in cosmopolitan areas), and light blue = Profile 5 (mostly women in precarious but educated environments). B. Spatial distribution of hospitalized senior TBI patient profiles in the canton of Geneva: multivariate typology and detection of significant risk zones using SPARR. The five colored point types represent the patient profiles identified through hierarchical agglomerative clustering (HAC): dark blue = Profile 1 (elderly women in nursing homes), blue = Profile 2 (isolated elderly men with short-term sequelae), light = Profile 3 (younger senior men, with rapid recovery), orange = Profile 4 (independent and affluent senior), and green = Profile 5 (Disadvantaged senior in nursing homes). The red areas indicate statistically significant relative risk clusters according to SPARR: light red for *p* < .050 and dark red for *p* < .010. An adaptive bandwidth was used for spatial risk estimation (significantly higher probability of TBI occurrence compared to the surrounding regions). Municipal boundaries and major waterways are also shown for geographic reference. Basemap from OpenStreetMap: https://www.openstreetmap.org

The SPARR analysis further highlighted specific high-risk areas for working-age and senior TBI patients. *For working-age*, one located in central Geneva, one near the French border (near the city of Presinge) and one in the countryside of the canton (near the city of Bernex). *For senior,* three located in central Geneva, aligning closely with the significant hotspots identified by the Join Count, and one at the French border near Dardagny and Avully. In this context, a significant *p*-value indicates the presence of a spatial risk cluster, meaning the probability of TBI occurrence is significantly higher (or lower) in the area compared to the surrounding regions. Overall, both methods converged in identifying similar high-risk zones, reinforcing the robustness and consistency of the spatial clustering results obtained through the Join Count and SPARR approaches (see Figure 3).

### Clustering

A hierarchical clustering analysis was conducted to identify distinct profiles of TBI patients across both the senior (60+) and working-age (15–59) populations, based on demographic, clinical, and contextual indicators. A total of ten clusters (5 for the senior and 5 for working-age) were identified, revealing diverse social and clinical trajectories associated with TBI (see Figure 4).

#### Clusters Specific to Seniors

- Senior Cluster 1: Institutionalized elderly women living in moderately advantaged, working-age neighborhoods (*n* = 191). High rates of EMS residency, moderate socioeconomic status.
- Senior Cluster 2: Isolated older men with long hospital stays and low home discharge rates (*n* = 171). Mostly living outside EMS facilities in non-senior neighborhoods.
- Senior Cluster 3: Younger senior men with short stays and rapid recovery (*n* = 115). High levels of autonomy and socioeconomic integration.
- Senior Cluster 4: Educated, affluent seniors from socioeconomically favorable environments (*n* = 73).
- Senior Cluster 5: Very elderly, dependent, and socioeconomically disadvantaged individuals (*n* = 53). High EMS residency and low levels of local activity.

#### Clusters Specific to the Working-age Population

- Working-age Cluster 1: Middle-aged men (40–59 years) with long hospitalizations, residing in overcrowded, low-senior neighborhoods (*n* = 94).
- Working-age Cluster 2: Young men (18–30 years), including all under 20s, representing typical early adulthood trauma cases with shorter stays (*n* = 83).
- Working-age Cluster 3: Adults aged 40–60 living in aging neighborhoods with high retiree concentrations and reduced economic dynamism (*n* = 71).
- Working-age Cluster 4: Men of all ages residing in cosmopolitan, working-age, and well-educated areas, concentrated in central Geneva (*n* = 44).
- Working-age Cluster 5: Women of various ages from relatively precarious but educated backgrounds, primarily located outside the urban core (*n* = 26).

### Spatial Distribution of working-age and senior clusters

Spatial mapping revealed clear geographic patterns across clusters (see Figure 5). Senior Clusters 1–3 were mainly concentrated in central Geneva, while Clusters 4 and 5 were more peripheral or institution-based. Among the working-age population, Cluster 4 was centered in Geneva’s city core, whereas Cluster 5 was more dispersed across suburban zones.

Cluster 2 (young adults) showed a relatively uniform distribution across the canton. Several areas (e.g., Versoix, Onex, Confignon, Eaux-Vives, and Veyrier) showed significant spatial clustering (*p* < .050), suggesting strong socio-spatial segmentation of TBI risk profiles.

These results highlight the diversity of TBI patient profiles and underline the role of age, gender, social context, and territory in shaping hospitalization pathways.

## Discussion

This study reveals how TBI are deeply shaped by age, social context, and geographic location of patients, producing different patterns of vulnerability and outcomes for seniors and the working-age population. Through spatial analysis, temporal exploration, and clustering methods, we identified profiles that are not only medically distinct but also territorially situated, reinforcing the need to understand TBI as both a clinical and societal phenomenon, for actionable prevention measures.

TBI risk clearly increases with age and gender, especially after 60 years and for men across the lifespan, corroborating previous observations ^14^. Among seniors, the burden is more than just numerical; it reflects a compounded vulnerability (physical fragility, cognitive decline, and social isolation) that converges to shape injury severity and outcomes. The observed decrease in home discharge rates with age illustrates the reduced recovery capacity or can lead to hypothesize an inadequate home environments or support systems for post-injury rehabilitation. Women, particularly those aged 70 and above, appear to experience more lasting hospitalizations, possibly due to greater longevity, lower physical and cognitive reserves, or less access to informal care networks ^15^. In contrast, among the working-age population, TBI is more often linked to contextual factors such as risk behaviors, occupational exposure, or dense urban environments ^16^, especially for men and can be associated to risky behaviors ^17^. While their overall recovery trajectories are more favorable, certain subgroups, especially middle-aged men experience prolonged hospital stays, indicating more complex trauma or slower recovery. These differences suggest that, for this group, injury context and socio-environmental exposure may matter as much as biological vulnerability. Spatial analysis revealed that TBI cases are not randomly distributed but tend to cluster in specific geographical zones, with patterns differing between working-age and senior populations.

Among seniors, clusters of TBI were observed in areas characterized by a combination of factors, including topographic risks (e.g., street slopes), a high density of nursing homes or elderly residents, and varying degrees of social vulnerability. This spatial clustering highlights how the features of the urban environment can amplify age-related risks (e.g., uneven pavements, poor lighting, or dense housing) which may turn into injury catalysts for already fragile individuals ^18^.

For the working-age population, spatial patterns were more fragmented but equally significant. Cluster analysis revealed diverse profiles: middle-aged men living in overcrowded areas, young men with minor injuries, and women from precarious but educated backgrounds. These findings show that social and spatial context shapes exposure and outcome, even when age-based physiological risk is lower. For instance, middle-aged men with long hospital stays were found in dense, working-age neighborhoods suggesting that injury severity may correlate with urban density, type of employment, or limited access to care ^19^. Moreover, and importantly, the spatial dynamics differed between age groups. Senior clusters were found in institution-rich or retirement-heavy areas, where frailty accumulates. In contrast, clusters in the working-age population reflect social mixing, residential stress, and behavioral risk, particularly in central or transitional urban zones. These patterns demonstrate that age and place interact in different way, depending on one’s life stage.

Concerning temporal patterns, a clear weekday trend was observed only among, whose TBI-related hospitalizations peaked at the beginning of the week. This may reflect weekly routines, such as increased movement after the weekend, for example a tendency of grocery shopping after the weekend in seniors, combined with reduced assistance on Mondays and Tuesdays ^20,21^. Another hypothesis is that, due to reduced medical resources and clinic availability during the weekend, additional events may be reported or diagnosed on Monday or Tuesday. This delay could contribute to more severe clinical outcomes and should be evaluated in future analyses. A seasonal effect was observed for both working-age and senior TBI patients, with significantly higher rates in autumn. Recent studies have observed an increased risk of road incidence ^22^ and TBI during autumn ^23^. A possible explanation is that autumn may present hidden dangers like wet leaves and rain, which may be underestimated compared to well-publicized winter risks like ice ^24,25^. In working-age adults, the absence of a weekday effect alongside a seasonal peak in autumn may indicate that TBI incidence in this group is less tied to predictable routines and more influenced by a wider range of social triggering contexts (work, sport, commuting).

The results of this study highlight the need for differentiated and localized prevention strategies. For seniors, efforts may focus on environmental adaptations in high-risk areas: improving sidewalk safety, increasing lighting, and deploying fall-prevention campaigns during high-risk seasons like autumn. Enhancing home care support and community follow-up is also essential for those living alone or outside institutional settings. In contrast, for the working-age population, prevention must take a more behavioral and occupational focus. Campaigns targeting middle-aged men in working-age neighborhoods, promoting workplace safety, or addressing commuting risks, could prove effective. Additionally, interventions should acknowledge the hidden vulnerability of certain groups, like women living in socially precarious contexts, whose risks are not always visible in traditional demographic indicators. Across both groups, urban design plays a critical role. Spatial clustering shows that certain neighborhoods concentrate both the risk and the resources to respond. This calls for urban health planning, where injury data inform public works, and where services like mobile care teams are directed to areas of need.

### Limitations

This study is limited by the absence of systematically recorded injury location and cause. Integrating these variables into hospital intake protocols would significantly enhance the power of spatial analyses. Future work will aim to link hospital data with real-time accident databases, urban risk indicators (e.g., noise, traffic, slope), and mobility data to better understand the interaction between people, place, and behavior. Distance to hospitals, transport access, and delays in care could also be included to model post-TBI outcomes and inequalities in emergency response.

## Conclusion

This study shows that TBI is not just the medical condition experienced by individuals. It is a territorial and social phenomenon shaped by the conditions and behaviors of urban life. Seniors and working-age adults are affected in different ways, at different times, and in different places. Yet in both groups, where people live, their social position, and the characteristics of their urban environment influence their risk of injuries and associated capabilities of recovery.

To be effective, TBI prevention must operate under a clear awareness of such complexity, with the need strategies targeted by age, geography, social profile, and season are essential. Urban planners, public health authorities, and healthcare providers should working-agely collaborate, using detailed injury data to create safer environments and build equitable systems of care. Ultimately, addressing the challenge and increasing epidemiological burden of TBI requires to see beyond the treatment. It requires anticipating injuries through spatial and social insight. In doing so, we can build cities and communities that not only heal but working-agely prevent injuries.

## Conflict of interest

The authors don’t report conflict of interests

## Funding

This research was supported by a grant from the THREESTONES Foundation, the European Association of Neurosurgery (EANS), and internal funds from the NeuroCentre of the Geneva University Hospitals (HUG). The funders had no role in data collection, analysis, interpretation, or the decision to publish the results.

## Acknowledgement

We would like to thank the members of the NeuroCentre of the University Hospitals of Geneva for their help with ethical submission and data storage.

## Notes

### Competing Interest Statement

The authors have declared no competing interest.

